# Bronchoscopy in COVID19 ARDS patients on mechanical ventilation – a prospective study

**DOI:** 10.1101/2021.02.02.21250362

**Authors:** Ravindra Mehta, Sameer Bansal, Ashwin Kumar, Anmol Thorbole, L Chakravarthi, Hariprasad Kalpakam

## Abstract

**Background:** Bronchoscopy has been done sparingly in COVID19 patients due to the risk of aerosol generation, with few reports describing its clinical utility. We describe a study on bronchoscopy in mechanically ventilated (MV) COVID-19 patients outlining the procedural, clinical, utilitarian and safety aspects.

**Methods:** Bedside bronchoscopy was performed in suspected or confirmed COVID-19 cases on MV; only positive cases were included in the study. Demographic, clinical, bronchoscopic and laboratory findings were noted and analysed.

**Results:** 98 procedures were performed on 61 patients, mean age of 62.1 years, 51 (83.6%) males. 42 patients (69%) had at least 1 co-morbidity. Major indications for bronchoscopy were new radiographic infiltrates with clinical deterioration, increased endotracheal tube (ETT) secretions and haemorrhagic secretions/hemoptysis. Common findings were copious secretions in 87 (88.8%), purulent in 61%, mucoid in 18%, haemorrhagic in 7% and frothy in 14% cases. Morphologically, hyperaemic airways were seen in 85 (86.7%) cases, ranging from mild (61%) to moderate-severe (39%). On the management front, antibiotics were changed in 31 (31.6%) cases based on bronchoscopic findings. Other significant changes included reduction or stopping of steroids and anticoagulation, fluid, and diuretic adjustment and ETT repositioning. The incidence of bacterial superinfection was also high (54% culture positivity for various bacteria), a significant number (94%) with multi-drug resistant organisms. Fungi were seen in 7 cases (7.1%). Pneumocystis jiroveci was not seen and cytology did not show any viral inclusions. Therapeutic mucus plug removal was done in 30 cases (30.6%), and hemoptysis control in 4% cases. The procedures were safe with no complications, and none of the HCW developed any COVID19 infection.

**Conclusion:** Bronchoscopy in critically ill MV COVID-19 patients contributes on both diagnostic and therapeutic fronts and can significantly influence management decisions. With adequate precautions and standard protocols, it is safe for both HCW and patients.

## Introduction

Bronchoscopy in COVID19 patients has been challenging due to the high aerosol-related risk to healthcare workers (HCW). (1,2) Several guidelines exist on bronchoscopic sampling in COVID19 patients. (3) Few reports define detailed clinical aspects, management impact and safety of ICU bronchoscopy in this patient population. This study on bronchoscopy in mechanically ventilated (MV) COVID ARDS (C-ARDS) patients was done in the peak of the COVID pandemic, and describes procedural, clinical, utilitarian and safety aspects of the procedure.

## Methods

Prospective observational study conducted at a tertiary committed COVID care center between August 25 and December 3, 2020. Approval for the study was granted by Institutional Ethics Committee Bio Medical Research Apollo Hospital, Bangalore. The study group included all MV ICU patients with initially proven or later confirmed COVID-19 who underwent bronchoscopy for clinical indications (Table 1). Bronchoscopy was deferred when any of the following were present; PEEP >10 cm H2O, hemodynamic instability, or operator’s perception of life-threatening deterioration during the procedure.

**Table 1:** Baseline characteristics of the patients included in the study.

| Baseline Characteristics |  |  |
| --- | --- | --- |
| No. of procedures | 98 |  |
| No. of patients | 61 |  |
| Mean age | 62.1 | SD ($\pm 11.5$) |
| Male sex | 51 | 83.6% |
| DM | 29 | 47.54% |
| HTN | 27 | 44.26% |
| CKD | 7 | 11.48% |
| CHD | 9 | 14.75% |
| Respiratory disease | 5 | 8.20% |
| Hypothyroidism | 2 | 3.28% |
| CVA | 2 | 3.28% |
| Malignancy | 2 | 3.28% |
| Median duration from symptom onset to hospitalization (Days) | 7 | IQR (4-10) |
| Median duration from symptom onset to MV (Days) | 10 | IQR (7-13.2) |
| Median duration from symptom onset to Bronchoscopy (Days) | 14 | IQR (10-20) |
| Median duration from MV to bronchoscopy (Days) | 2.5 | IQR (1-6.5) |
| Indications for Bronchoscopy |  |  |
| New Radiological Infiltrates | 70 | 71.43% |
| Segmental collapse | 27 | 27.55% |
| Increased ET secretions | 36 | 36.73% |
| Hemoptysis/Bloody secretions in ET | 3 | 3.06% |
| Tracheostomy | 7 | 7.14% |

The following variables were recorded: Demographic and clinical parameters such as age, gender, duration of symptoms prior to hospitalization, presence of co-morbidities [diabetes, hypertension, chronic kidney disease (CKD), ischemic heart disease (IHD)], and duration of ventilatory support prior to procedure. Procedure details included indications, findings, relevant microbiological and cytological tests, and management changes following bronchoscopy. Safety aspects from both the patient and the HCW perspective were also studied.

### Procedure

Bronchoscopy after informed consent was performed by 3 different operators. A bronchoscopy technician, a respiratory therapist and an ICU nurse were present for every procedure. All health care workers (HCW) used adequate personal protective equipment PPE (P-100 respirator, impermeable coverall, face shield and double layered gloves). All HCW’s self-monitored for symptoms for the duration of the study and if symptomatic, a nasopharyngeal swab was taken for COVID-19 RT-PCR.

The procedure was performed at the bedside in the ICU, which had > 20 air exchanges/hour. Negative pressure isolation rooms were not available. Sedation included midazolam and fentanyl and short-acting neuromuscular blockade with atracurium to prevent any aerosol generating cough. Pre-procedure, FiO2 was increased to 100% for 20 mins. During the bronchoscopy, the ventilator was transiently disconnected for periods less than 1 minute to minimize aerosol generation. A rapid bronchoscopy was done, with close monitoring of SpO2 and vital parameters, with brief in-and-out runs with the bronchoscope as needed. As a safety measure, patients in prone position were maintained in the same position to mitigate procedure related desaturation.

Pooled washings (average 80-100 ml from multiple segments) were done in view of the need for multi-segment sampling and concern of desaturation with a larger volume BAL. Samples were collected and analysed for laboratory investigations including the COVID-RT PCR.

### Statistics

Data was tabulated and analysed using SPSS (ver. 25.0, SPSS Inc). Results were analysed in a descriptive fashion as number and percentages, mean and standard deviation, median and inter quartile range (IQR).

## Results

98 procedures were done in 61 MV C-ARDS patients. 41 patients had one bronchoscopy procedure, while 20 patients had repeat procedures, for various indications (Table 1).

### A. Demographics

Baseline characteristics of the study group including demographic and co-morbidity details are mentioned in Table 1. Of note, 69% patients (42/61) had at least 1 comorbidity, while 3 patients (5%) had a combination of DM with chronic respiratory illness, and 6 patients (10%) had DM with CKD.

### B. Timelines

Median duration from symptom onset to hospitalization was 7 days (IQR; 4-10), symptom onset to MV was 10 days (IQR; 7 – 13.2), symptom onset to bronchoscopy was 14 days (IQR; 10-20), MV to bronchoscopy was 2.5 days (IQR; 1-6.5).

### C. Indications and findings

Common indications included clinical worsening with new or increasing infiltrates on the chest radiograph (CXR) in 70 (71.4%), segmental collapse on CXR in 27 (27.6%), increased endotracheal (ETT) secretions in 36 (36.7%) and hemoptysis in 3 (3.1%) cases. Copious increased ETT secretions necessitated repeat procedures. 1 patient had near complete ETT block due to thick inspissated secretions. 7 patients underwent bronchoscopy during tracheostomy as a combined strategy to facilitate a quick procedure and perform airway evaluation and sampling. (Table 2).

**Table 2:** Major bronchoscopic findings.

| Bronchoscopy Findings |  | N | % |
| --- | --- | --- | --- |
| **Increased secretions** |  | **87** | **88.78%** |
|  | Purulent | 53 | 60.92% |
|  | Clear (mucoid) | 16 | 18.39% |
|  | Frothy | 12 | 13.79% |
|  | Bloody | 6 | 6.90% |
| **Frank haemorrhage** |  | **4** | **4.08%** |
| **Inflamed/Hyperraemic airways** |  | **85** | **86.73%** |
|  | Mild | 52 | 61% |
|  | Moderate to severe | 33 | 39% |
| **Mucus plugging** |  | **30** | **30.61%** |
| **Others** |  |  |  |
|  | Tracheomalacia | 7 | 7.14% |
|  | Bronchiectasis | 5 | 5.10% |
|  | Mucosal ulceration | 2 | 2.04% |
|  | Incidental mass LUL | 1 | 1.02% |

The commonest bronchoscopic findings were increased secretions, seen in 87 (88.8%) cases. 53 (61%) had thick purulent secretions, 16 (18.4%) had clear mucoid secretions, 12 (14%) had frothy secretions, and 6 (7%) had haemorrhagic secretions. Airway hyperaemia was seen in 85 cases (87%), of which 52 (61%) was mild, and 33 (39%) had moderate to severe hyperaemia. Mucus plugging was seen in 30 (30.6%) cases, which improved after therapeutic suctioning. Mild bleeding was noted in 4 cases (4.1%). Other findings included suspected bronchiectasis in 5 patients evidenced by easy passage of the scope beyond the 5th generation bronchus, tracheomalacia in 7 patients, mucosal ulceration in 2 patients, and an incidental polypoidal mass lesion (malignancy) in 1 patient. (Table 2) (Fig.1)

**Fig 1:**
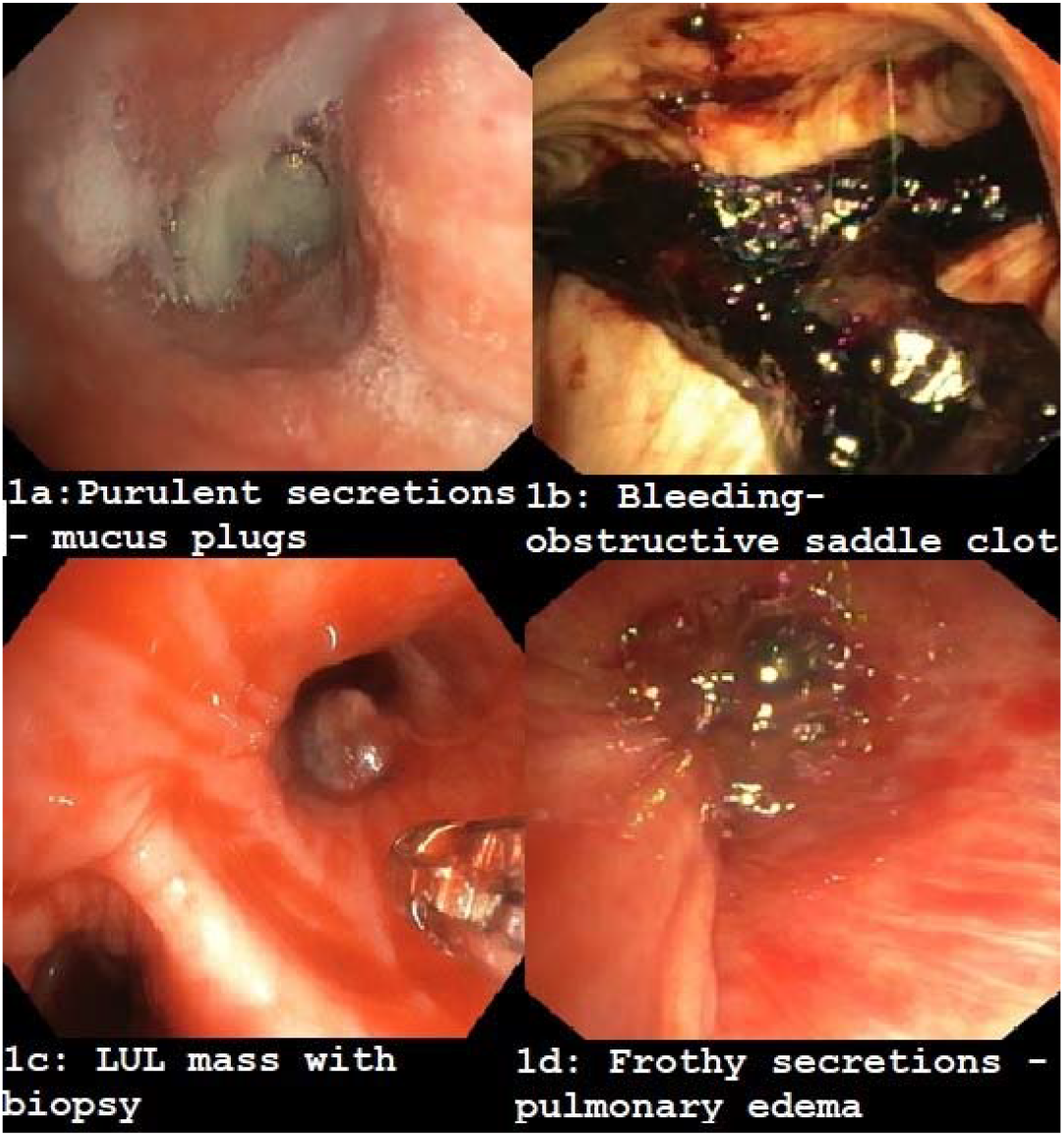
Various bronchoscopic findings. 1a: Purulent secretions with obstructive mucus plug. 1b: Endobronchial bleeding – obstructive saddle carinal clot seen 1c: Incidentally detected LUL mass – biopsy confirmed atypical carcinoid. 1d: Frothy secretions due to pulmonary edema

### D. Microbiology

The bacterial microbiology showed the following: Gram’s stain showed pus cells in 79 cases (80.6%), gram-positive organisms in 2 (2.1%) and gram-negative bacilli in 34 cases (34.6%). The final culture was sterile in 45 (46%), while it was positive in 53 cases (54%). The most common organisms isolated were *Klebsiella pneumoniae* in 29 (29.5%), *Acinetobacter baumanii* in 8 (8.3%), *Burkholderia cepacia* in 4 (4.2%), *Enterobacter cloacae* in 3 (3.1%) and *Acinetobacter iwofii, Providencia stuartii* & *Serratia marcesans* in 2 cases each. *MRSA, Pseudomonas aeruginosa, Morganella morganii, Stenotrophomonas maltophilia* & *Citrobacter freundi* were cultured in 1 patient each. (Table 4). 3 patients grew more than 1 bacteria. 94.3% (50/53) of these organisms were multidrug resistant (MDR).

**Table 3:**
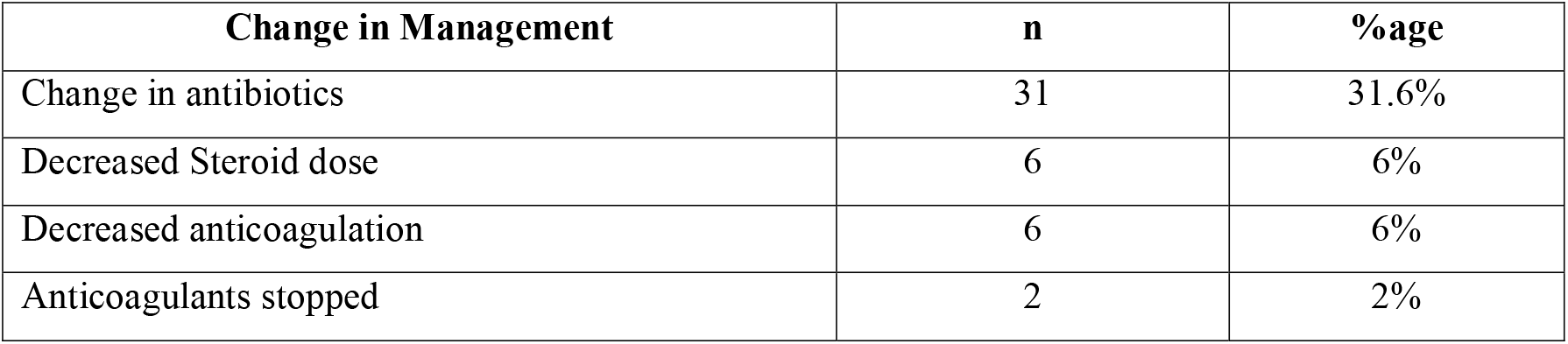
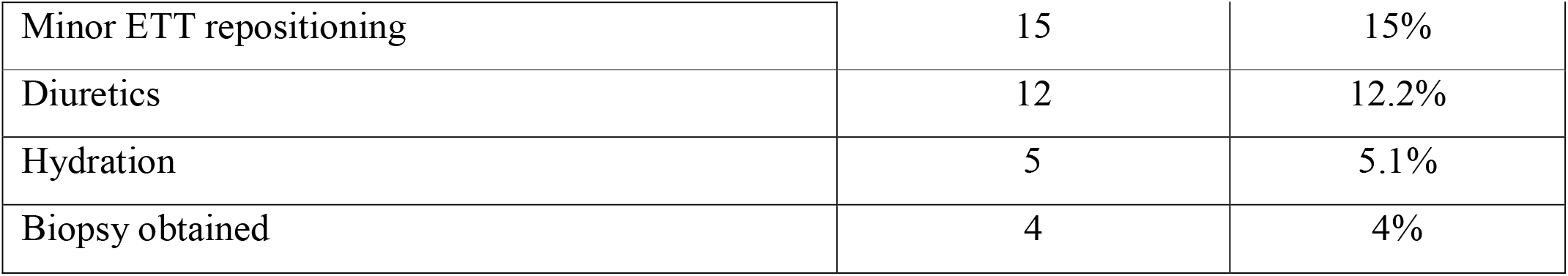
Change in management following bronchoscopy.

| Change in Management | n | %age |
| --- | --- | --- |
| Change in antibiotics | 31 | 31.6% |
| Decreased Steroid dose | 6 | 6% |
| Decreased anticoagulation | 6 | 6% |
| Anticoagulants stopped | 2 | 2% |
| Minor ETT repositioning | 15 | 15% |
| Diuretics | 12 | 12.2% |
| Hydration | 5 | 5.1% |
| Biopsy obtained | 4 | 4% |

**Table 4:** Bacterial culture results.

| Culture positivity | N | % |
| --- | --- | --- |
|  | **53** | **54.08** |
| *Klebsiella pneumoniae* | 29 | 54.72 |
| *Acinetobacter baumannii* | 8 | 15.09 |
| *Burkholderia cepacia* | 4 | 7.55 |
| *Enterobacter cloacae* | 3 | 5.66 |
| *Acinetobacter iwoffi* | 2 | 3.77 |
| *Providencia stuartii* | 2 | 3.77 |
| *Serratia marcesans* | 2 | 3.77 |
| *Citrobacter freundii* | 1 | 1.89 |
| *MRSA* | 1 | 1.89 |
| *Pseudomonas aeruginosa* | 1 | 1.89 |
| *Morganella morganii* | 1 | 1.89 |
| *Stenotrophomonas maltophilia* | 1 | 1.89 |

Fungal evaluation in the samples showed the following: 7 patients had a positive KOH mount, of which 5 showed budding yeast with septate hyphae, while 2 had presence of aseptate hyphae. Fungal culture was positive in 2 patients. BAL galactomannan was sent in 4 patients and was elevated in all 4. Appropriate anti-fungal agents were added in all the cases.

Silver stain did not show *Pneumocystis jiroveci* and all samples were negative for acid-fast bacilli. In 1 patient, COVID diagnosis was confirmed on washings RT-PCR, after 2 NP swabs were negative.

### E. Cytology and Histopathology

The cytology showed no evidence of viral inclusions or any other significant abnormality. Histopathologically, biopsies done on 4 abnormal endobronchial lesions showed non-specific inflammation, and the LUL mass biopsy showed atypical carcinoid tumor.

### F. Impact of bronchoscopy

In C-ARDS, clinical worsening with new/increasing infiltrates had several possibilities including primary disease progression, superinfection (bacterial, fungal, other) and non-infectious causes (fluid shifts, atelectasis, pulmonary infarction). There was scant literature to guide management at this point in the pandemic, and lower respiratory tract sampling was limited with a restricted suctioning policy. With clinical deterioration, there was a need to supplement clinical judgement with objective data, which was provided by bronchoscopy. Bronchoscopy impacted management in the following ways (Table 3):

1. Antibiotics were changed/escalated in 31 (31.6%) cases immediately based on bronchoscopic findings of copious purulent secretions. These were not seen in the ETT prior to bronchoscopy in 40/53 (75.5%) patients with purulent secretions and was a new finding. Since most of these patients (94%) grew multi-drug resistant (MDR) organisms sensitive only to the polymyxin group of antibiotics, we changed our policy to empiric polymyxin antibiotics for any patients suspected of infection on MV.
2. When extensive purulent secretions were seen, corticosteroid dosage was reduced/stopped.
3. Anticoagulation was reduced from intermediate to preventive (equivalent of enoxaparin 40mg twice a day to 40mg once a day) in 6 patients (6%) with haemorrhagic secretions, while it was completely stopped for 2 patients with significant persistent ooze. Iced saline with diluted adrenaline (1:10,000) was used to achieve haemostasis.
4. ETT repositioning done in 15 patients (15.3%) due to close proximity to carina (< 1 cm) especially with a prone-supine protocol in place, considering the possibility of caudal bronchial displacement.
5. Fluid administration was reduced, and diuretics added in 12 patients (12.2%) based on the visual perception of pulmonary edema (frothy copious upwelling secretions).
6. Therapeutic suctioning of thick mucus plugs was done in 30 (30.6%) cases, of which 14 (14.2%) had very thick inspissated obstructive secretions.

Assessing risk to the HCW’s, none of the HCWs developed any symptoms or COVID positivity during the study period.

## Discussion

Our study of bronchoscopy in MV COVID-19 critically ill patients describes diagnostic and therapeutic aspects, including morphological details, microbiological and pathological aspects, and procedural and safety aspects.

Bronchoscopy helped obtain more information to guide management when there was limited information in a new pandemic. A fundamental limitation in the MV-CARDS patients during the pandemic was restricted suctioning due to aerosol risk, limiting many aspects of diagnosis and ventilator management. (1) Bronchoscopic inspection led to immediate changes including change of antibiotics, reduction of immunosuppression and anticoagulation, modification of the fluid strategy, ETT tube adjustment and detection of unexpected findings, such as a malignant mass (Table 2). Microbiologically, the Gram’s stain and subsequent culture reports were used to adjust antibiotics in line with standard principles. Another important result was proving COVID positivity in washings when earlier NP swabs were negative.

Few studies have been published on bronchoscopy in severe COVID-19 patients. Torrego et al performed 101 bronchoscopies in 93 COVID-19 patients early in the pandemic. (4) The median duration from MV to procedure was 6.6 days (range 1-17). Bruyneel et al. performed 90 bronchoscopies in 32 ICU patients between 6 March and 21 April 2020. (5) Baron et al. performed 28 bronchoscopies between March 31 and June 2020 on 24 COVID-19 patients. (6) The median time [IQR] from MV to BAL was 16 [10-21] days. In our study, median symptom-onset (SO) to hospitalization duration was 7 (IQR;4-10) days, SO to MV was 10 days (7 – 13.2) and SO to bronchoscopy was 14 days (10-20), while MV to bronchoscopy was 2.5 days (1-6.5). The timing of bronchoscopy in our study was based on clinical indications, with the observation that longer duration of illness/MV and use of immunosuppressive therapy (uniform steroids and occasional tocilizumab) was a risk factor for MDR infection. Salient comparison points from these studies are highlighted in Table 5.

**Table 5:** Major COVID bronchoscopy studies.

|  | **Torrego et al.** | **Bruyneel et al.** | **Baron et. al.** | **Mehta et. al.** |
| --- | --- | --- | --- | --- |
| **No. of procedures** | 101 | 90 | 28 | 98 |
| **No. of patients** | 93 | 32 | 24 | 61 |
| **Median duration from MV to bronchoscopy** | 6.6 days | NA | 16 days | 2.5 days |
| **Major indications** | - Radiological infiltrates<br>- Increased secretions | - Unexplained hypoxemia<br>- Microbiological sampling<br>- Mucus plug removal | - Microbiological sampling | - New radiological infiltrates<br>- Increased secretions<br>- Hemoptysis |
| **Major findings** | - Increased secretions<br>- Mucus plugs<br>- Normal to mild hyperaemia | - Mucus plugs | NA | - Increased secretions<br>- Mucus plugs<br>- Hyperaemic airway in 85% cases |
| **Bacterial superinfection** | 28.6% | 60% | 86% | 54% |
| **Most common microorganism** | 1. Pseudomonas aeruginosa<br>2. Staphylococcus aureus |  | NA | 1. Klebsiella Pneumonia<br>2. Acinetobacter baumannii |
| **Fungal** | NA | 16 samples (17.7%) | 25% cases | 7 cases (7.1%) |

Our study bears similarities and also contrasts with prior COVID19 bronchoscopy reports. Our main indications for bronchoscopy were increased secretions, new or increasing radiographic infiltrates, segmental collapse, suspicion of ETT blockage or copious secretions causing desaturation, and haemorrhagic ETT secretions. This experience is in part similar to the study by Torrego et. al. where major indications for bronchoscopy were radiological and/or clinical deterioration suggesting possible superinfection (63/101), and airway secretion management with/without atelectasis (38/101). (4) Bruyneel et al. performed bronchoscopies for unexplained worsening of hypoxemia and microbiological sampling to guide appropriate antimicrobials or remove bronchial mucous plugs. (5) Baron et al did BAL for a microbiological purpose in all cases -to confirm SARS-CoV-2 infection (7%), after a negative RT-PCR NP swab, for suspicion of ventilator-associated pneumonia (39%) or invasive aspergillosis (14%) or to rule out superinfection before starting a corticosteroid course (43%). (6) All procedures were done in MV patients, similar to our study.

The most common finding in our study was increased secretions seen in 87 (88.8%) cases, which included thick purulent secretions in 53 (61%), clear mucoid secretions in 16 (18.4%), frothy secretions in 12 (13.8%), and bloody secretions in 6 (7%) patients. Torrego et. al. reported diffuse, white, and jelly-like secretions, difficult to suction in 95% patients. (4) The therapeutic aspect of the procedure also has been highlighted in literature. In the above series, in 12 cases, muco-hematic plugs were removed using saline and a mucolytic agent. Bruyneel et al. report that purulent plugs were removed in 33 procedures (37% cases). They further report that “in the majority of these patients, very thick and dry plugs (like limestone) were stuck in the ETT. The tube became quickly dirty and needed to be replaced more often than usual”. (5) We had different observations-therapeutic suctioning of thick mucus plugs were done in 30 (30.6%), and only 14 (14.2%) of these had very thick inspissated obstructive secretions. Though we had 1 case where the ETT was almost completely blocked and had to be replaced, we did not commonly face this due to a robust humidification strategy and a strategy of early bronchoscopy if secretion related issues were suspected. With limited suctioning in the active disease phase, the dual role of bronchoscopy as a lower respiratory tract sampling modality and airway de-obstruction therapeutic tool was invaluable.

Another interesting observation in our study was hyperaemic and inflamed bronchial mucosa in 86.7% cases. This could be attributable to COVID inflammation in general, viral, or bacterial infection, or routine use of prophylactic/ intermediate anticoagulation. Torrego et. al. reported normal or mildly hyperaemic bronchial mucosa in most patients. (4)

There was bacterial superinfection in 54% cases, proven by cultures with significant colony counts (> 106/ml). Torrego et al reported 28.6%, Bruyneel et al 60% and Baron et al 86% positive culture cases. (4-6) The spectrum of microorganisms varies -in previous studies, the commonest microorganisms isolated were *Pseudomonas* and *Staphylococcus aureus*, while our study had predominantly *Klebsiella pneumonia* and *Acinetobacter baumanii*. This variation may not be specific to COVID-19 but likely represents superinfection with resident ICU flora in sick patients with comorbidities, prolonged ICU stay, and a viral pneumonia complicated with uniform use of steroids and antibiotics. The utility of BAL was shown by Baron et al. for detection of super-infection. Compared to other less invasive microbiological tests, BAL identified at least one previously undetected pathogen in 46% cases. (6)

We had fungal smear positivity in 7 cases (7%), BAL galactomannan positive in 4 cases and fungal culture positive in 2 cases. Bruyneel et al report fungi in 16 samples, but all were culture/galactomannan negative. (5) Baron et al. reported Aspergillus spp. isolation by culture/PCR in 7 (25%) cases. (6) Case series by Koehler et al. and van Arkel et al. also suggest a high incidence of Aspergillosis −20-25% in critically ill COVID-19 patients. (7,8) We did not find such a high incidence of fungal infection in our patients despite comorbidities, broad spectrum antibiotics and steroids. Additionally, there was no infection with P. jiroveci noted in the entire cohort. In terms of confirming COVID19, 1 patient had the diagnosis confirmed on fluid RT-PCR after 2 negative NP swabs. In the study by Baron et. al., COVID-19 diagnosis was confirmed on BAL RT-PCR in 2 patients (7%). (6) Patrucco et al reported that of the 120 patients with 2 negative swabs, SARS-CoV-2 was isolated (27.5%) on BAL. (9)

Bronchoscopy in these critically ill C-ARDS patients had a significant impact on management, both immediate and short-term as mentioned earlier. In this regard, our findings are consistent with the findings of prior studies and extend and support the role of bronchoscopy in this patient population for more precise decision-making and management changes. Torrego et al. based on BAL introduced a new antibiotic in 15/18 (83%) patients. (4) Bruyneel et al. state that bronchoscopy led to antibiotic adaptation in 18% of total and 31% of positive microbiological samples, a benefit clinically relevant to patient management. (5) Baron et al. mentioned that BAL impacted decision making in 71% cases: introduction, continuation, switch, or withdrawal of antimicrobial therapy in 50% cases, and decision to start (21%), or not (21%) corticosteroid therapy. (6) In our study, in 31.6% cases, antibiotics were escalated based on copious purulent bronchial secretions, with subsequent confirmation on culture. The change in antibiotic policy, with empirical polymyxins being used with suspicion of infection, was also part of a course correction as we analysed our preliminary culture results.

Other important decisions coinciding with antibiotic escalation were to de-escalate/stop corticosteroids when copious purulent secretions were noted, as a systematic immunosuppression reduction strategy. Anticoagulants were reduced when the mucosa was too hyperaemic and bleeding on touch and discontinued in cases of visibly bloody secretions or a persistent ooze, especially in patients with hemoptysis. In terms of fluid management, fluids were reduced, and diuretics introduced in patients with evident pulmonary edema (“frothy secretions”) (12%). Hydration and humidification were enhanced with a visibly dry mucosa and inspissated secretions in 5 patients. These findings and consequent management adjustments were possible only with bronchoscopic evaluation.

Histopathological evaluation of an incidental mass noted showed atypical carcinoid and the 4 other bronchial biopsies of abnormally inflamed areas showed non-specific inflammation. There were no complications in the biopsy process. For hemoptysis and haemorrhagic secretion management, bleeding control was done with local measures, combined with reduction/cessation of anticoagulation in 8 patients.

Our procedure technique factored in both patient and HCW safety and was like that reported earlier, namely pre-oxygenation with 100% FiO2, a quick procedure and scope removal followed by reinsertion in case of desaturation. We additionally interrupted ventilation for brief periods while performing bronchoscopy to minimize aerosol generation. (10) Bronchoscopy in COVID-19 patients when done with adequate precautions is relatively safe with low risk to HCWs. This fact is reinforced in various studies (11,12), with only Torrego et al. reporting 1 operator developing COVID-19 infection. (4) In our study, none of the HCW’s developed COVID19 infection, due to appropriate PPE, adequate ICU air exchanges, an additional P-100 respirator conferring add-on protection, combined with brief disconnection of the ventilator for short periods. In terms of Patient safety measures included maintaining prone position for bronchoscopy whenever possible and using washings instead of BAL in hypoxemic patients.

Unique aspects of our study include the value of additional information obtained from bronchoscopy that influenced clinical management, especially in patients where there were no ETT secretions, and limited suctioning as per protocol. Numerous management changes, and antibiotic policy changes done midway were based on information from the procedure. The technique and enhanced PPE (3M respirator with P100 filter) ensured safety both for the patient and the HCW despite the absence of negative pressure. This is one of the few studies with uniform steroid use in C-ARDS, and bronchoscopy helped to define both the microbiological impact of a uniform steroid strategy as well as the decision to continue steroids. In addition, we looked for and did not see any additional mucormycosis, tuberculosis or PJP.

Our study has certain limitations – it is restricted to critically ill MV patients at various durations of C-ARDS and represents the sickest end of the spectrum. We could not do molecular studies and were not able to do galactomannan in all the samples. Our sample is typically pooled washings from multiple areas with net volumes 80-100 ml, as these patients were significantly hypoxemic, and a larger volume BAL was perceived as a potential unsafe risk in this critically ill MV cohort. We preferred bronchoscopy to mini-BAL due to the expanded diagnostic and therapeutic role of conventional bronchoscopy as mentioned above.

## Conclusion

Our study of bronchoscopy in critically ill MV COVID19 patients is one of the few reports which describes the utility and safety of bronchoscopy in this cohort at the peak of the pandemic. Important morphological, microbiological, and pathological data was obtained with reasonable safety for both HCW’s and patients. Bronchoscopic intervention was valuable on diagnostic, therapeutic and management altering fronts. It should be strongly considered as a useful and safe modality to enhance effective treatment of these critically ill patients at the point of unexplained clinical deterioration.

## Data Availability

All the data pertaining to this study is available for review as required.

## Acknowledgments

We like to thank Dr. Michael Cutaia and Dr. Mohammed Munavvar for reviewing the manuscript and providing inputs.

